# The Mammillary Body-Fornix Gate in Long COVID An exploratory structural and diffusion MRI study of tremor-like symptoms, internal vibrations, and neuromuscular fatigue

**DOI:** 10.64898/2026.07.31.26359395

**Authors:** Christof Peter Ziaja, Susanne Yvette Young, Michael Stark Sadre-Chirazi, Grzegorz Zurék, Jan Sedlacik, Felicia-Marie Wright

## Abstract

**Background:** Long COVID frequently includes post-exertional malaise, neuromuscular fatigue, internal vibrations, tremor-like symptoms, orthostatic intolerance, sleep disturbance, cognitive dysfunction, and autonomic instability. Neuroimaging studies in Long COVID and post-infectious ME/CFS have identified abnormalities across limbic, thalamic, mesiotemporal, brainstem, cerebellar, and white-matter systems, but no single established circuit accounts for this clinical phenotype. We examined whether abnormalities cluster at the mammillary body-fornix-superior tuberal hypothalamic interface and within connected brainstem-cerebellar pathways.

**Methods:** Structural MRI and diffusion tensor imaging were analysed in 88 participants with clinician-diagnosed Long COVID and 34 healthy controls (total N = 122). Thirty-three patients were classified as bedridden. Volumetric and diffusion analyses focused on the mammillary bodies, superior tuberal hypothalamic region, fornix, dorsal and median raphe, midbrain reticular formation, and cerebellar peduncles. Fractional anisotropy was used as an index of white-matter microstructural organisation. Exploratory clinical associations included motor impairment, proprioceptive dysfunction, autonomic symptoms, fatigue, internal vibrations, and tremor-like symptoms.

**Results:** Three mammillary-body volume phenotypes were identified: reduced, enlarged, and control-range. Left and right mammillary-body volumes differed across groups, with large effect estimates. Segmentation showed narrowing or loss of a visible internal passage at the superior tuberal-mammillary interface, while the fornix showed altered diffusion measures and reduced tract coherence in the hypothesised gate region. Additional findings included lower superior cerebellar peduncle volume, lower middle cerebellar peduncle fractional anisotropy, and lower dorsal raphe and midbrain reticular formation volumes in Long COVID.

**Conclusions:** The findings support an exploratory mammillary body-fornix gate model in which a vulnerable periventricular hypothalamic-limbic interface may contribute to network dysfunction in a subgroup of patients with severe Long COVID. The data do not establish direct viral invasion, a coronavirus entry route, axonal destruction, or a single causal pathway. Prospective replication with standardised acquisition, preregistered regions of interest, correction for multiple comparisons, objective movement and autonomic measures, and longitudinal follow-up is required.

**Ethics application 2022-100867-BO-ff:**

1. Ethics approval was granted by the Ethics Committee of the Hamburg Medical Association, Germany, on September 5, 2022, under the title; MRI Biomarkers in Chronic Fatigue, by Prof. Dr. Rolf Stahl.
2. The project complies with the ethical and professional requirements. The Ethics Committee approves the project. The Ethics Committee operates on the basis of German law and professional regulations, as well as in accordance with ICH-GCP.

## 1. Introduction

Long COVID is a chronic and frequently disabling post-infectious condition characterised by fatigue, post-exertional malaise (PEM), cognitive dysfunction, dysautonomia, sleep disturbance, pain, exertional intolerance, and multisystem symptoms (Al-Aly et al., 2024; Davis et al., 2023; Peluso & Deeks, 2024). The clinical phenotype overlaps substantially with post-infectious myalgic encephalomyelitis/chronic fatigue syndrome (ME/CFS), particularly where PEM, impaired function, unrefreshing sleep, cognitive symptoms, and orthostatic intolerance are prominent (Walitt et al., 2024). Current evidence supports interacting biological mechanisms, including immune dysregulation, complement activation, endothelial and microvascular dysfunction, autonomic impairment, altered metabolism, barrier dysfunction, and central nervous system involvement. These mechanisms are heterogeneous and should not be collapsed into a single explanatory model.

Neurological and autonomic manifestations are common in Long COVID. Neuroimaging studies have reported changes in limbic and mesiotemporal tracts, thalamic and brainstem metabolism, brainstem microstructure, choroid plexus volume, blood-brain barrier integrity, and broader subcortical network dynamics (Diez-Cirarda et al., 2025; Douaud et al., 2022; Greene et al., 2024; Guedj et al., 2021; Rua et al., 2024). In post-infectious ME/CFS, PET, diffusion MRI, and functional connectivity studies have likewise implicated the thalamus, hippocampus, amygdala, midbrain, and intra-brainstem networks (Barnden et al., 2019; Nakatomi et al., 2014; Shan et al., 2023). This literature supports investigation of anatomically related regions, but it does not establish a single formally defined brainstem-hypothalamic-limbic-thalamic circuit or a directional viral entry pathway.

The present study focuses on a specific anatomical convergence zone: the mammillary body-fornix-superior tuberal hypothalamic interface. The fornix carries major hippocampal and subicular outputs to the mammillary bodies and other diencephalic targets. The mammillary bodies, in turn, are embedded within the posterior hypothalamic region and participate in extended hippocampal-diencephalic memory networks (Aggleton et al., 2010, 2022; Bubb et al., 2017; Vann & Nelson, 2015). Adjacent superior tuberal and paraventricular hypothalamic regions contribute to endocrine, autonomic, metabolic, thermoregulatory, sleep-wake, and immune-to-brain regulation (Herman et al., 2016; Saper & Lowell, 2014).

We use the term mammillary body-fornix gate as an operational, testable model rather than a recognised named circuit. The gate refers to the structural and functional interface through which forniceal fibres approach the mammillary bodies within a small periventricular hypothalamic region. We hypothesised that morphological and diffusion abnormalities at this interface, together with abnormalities in connected brainstem-cerebellar systems, may be associated with internal vibrations, tremor-like symptoms, neuromuscular fatigue, autonomic instability, and cognitive symptoms in a subgroup of patients with severe Long COVID.

## 2. Neuroanatomical and pathophysiological rationale

### 2.1 Mammillary body, fornix, and superior tuberal hypothalamus

The fornix is a major white-matter output pathway of hippocampal formation. Postcommissural fibres arise prominently from the subiculum and project to the mammillary bodies, while mammillary projections reach the anterior thalamic nuclei through the mammillothalamic tract. These structures contribute to extended hippocampal-diencephalic networks involved in memory, spatial context, and behavioural orientation (Aggleton et al., 2010, 2022; Bubb et al., 2017). The classical serial Papez circuit is now considered an oversimplification; accordingly, the present paper describes interconnected pathways rather than a single closed circuit.

**Figure 1.**
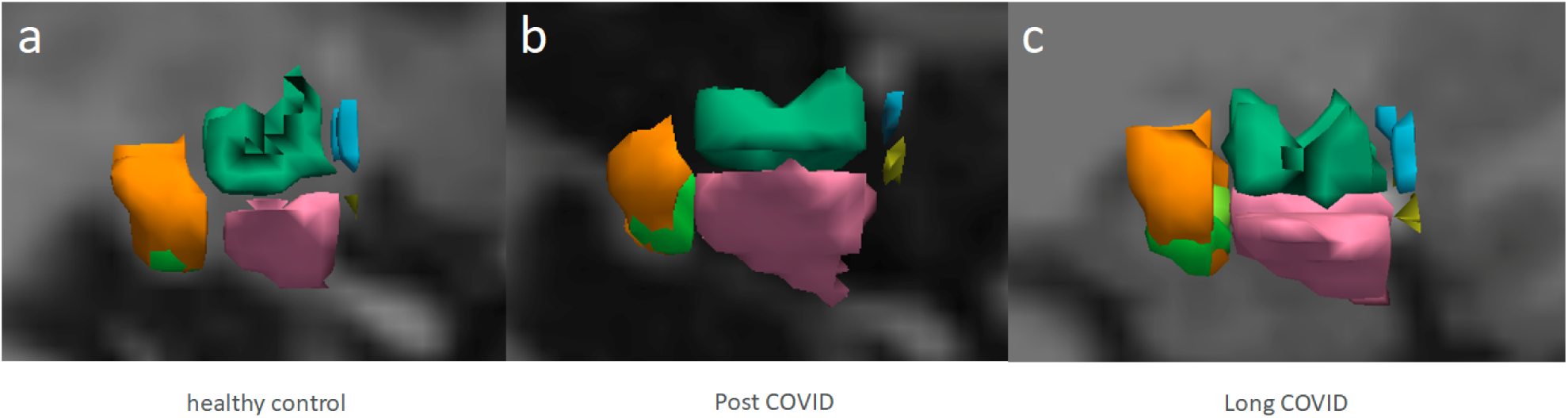
Sagittal examples of hypothalamic segmentation in a healthy control (a), a post-COVID participant (b), and a Long COVID participant (c), generated in FreeView using the FreeSurferColorLUT. Coloured regions show the right anterior-superior (blue), anterior-inferior (yellow), inferior tuberal (pink), superior tuberal (green), posterior hypothalamic region (orange), and mammillary body (light green). In panels b and c, reduced visible separation is observed between the inferior tuberal and posterior hypothalamic regions.

The mammillary bodies are positioned within the posterior hypothalamus, close to the third ventricle and adjacent hypothalamic regions involved in autonomic and endocrine control. The superior tuberal and paraventricular region participates in hypothalamic-pituitary-adrenal regulation, sympathetic outflow, osmotic and metabolic signalling, immune-to-brain communication, and behavioural-state coordination (Herman et al., 2016; Saper & Lowell, 2014). Changes in this small periventricular region could therefore affect multiple functions without implying that one structure alone explains the Long COVID phenotype.

**Figure 2.**
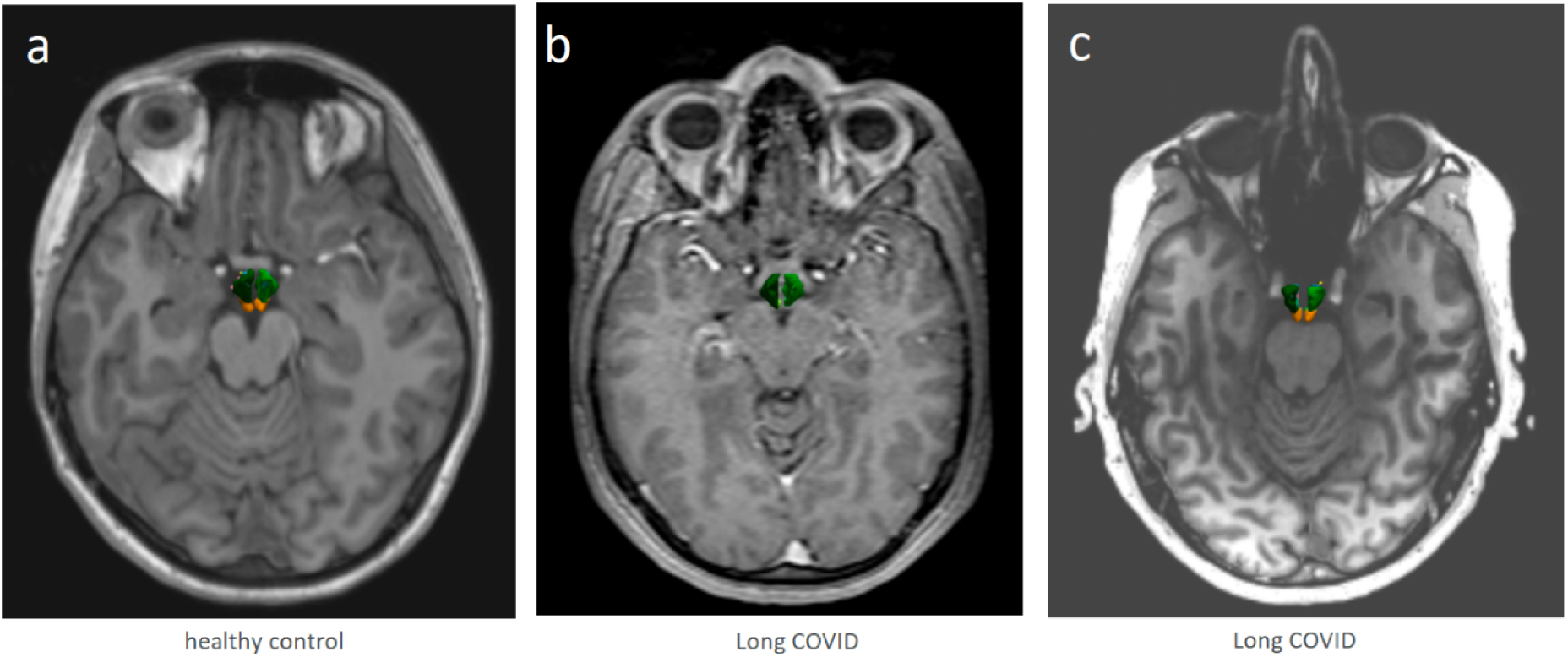
Axial 3D isosurface views of hypothalamic segmentations. Panel a shows a 3-T scan acquired using the Siemens Skyra protocol used for the study cohort; panels b and c illustrate comparison segmentations from 1-T and 7-T scans. Across the illustrated Long COVID examples, segmentation showed narrowing or loss of a visible internal passage in the superior tuberal/periventricular region.

In this study, the visually described internal openings were operationalised as segmentation-defined narrowing or loss of a visible internal passage in the superior tuberal/periventricular region. This is a morphological observation and does not, by itself, demonstrate mechanical compression, axonal loss, inflammation, or obstruction of cerebrospinal-fluid flow.

### 2.2 Tremor-like symptoms, internal vibrations, and brainstem-cerebellar regulation

Internal vibrations and tremor-like symptoms are phenomenological descriptions rather than established movement-disorder diagnoses. They may arise through multiple mechanisms, including altered autonomic state regulation, motor-network gain, sensory processing, brainstem arousal, or peripheral neuromuscular processes. Thermoregulatory shivering is a distinct physiological response involving thermal sensing, hypothalamic integration, brainstem premotor pathways, sympathetic activation, and skeletal-muscle thermogenesis (Tan & Knight, 2018); it should therefore only be used when the observed episodes are compatible with that mechanism. The present model treats the reported movement symptoms as a phenotype requiring objective characterisation rather than as evidence of a specific lesion or thermoregulatory disturbance.

The dorsal and median raphe, locus coeruleus, midbrain reticular formation, periaqueductal grey, pontine and medullary nuclei, and cerebellar peduncles contribute to arousal, sleep-wake transitions, pain modulation, autonomic tone, postural control, and sensorimotor integration. Prior ME/CFS and Long COVID studies have reported altered intra-brainstem connectivity, brainstem inflammatory signatures, and brainstem imaging abnormalities (Baraniuk et al., 2022; Barnden et al., 2019; Matschke et al., 2020; Radke et al., 2024; Rua et al., 2024). These observations provide a plausible context for the brainstem-cerebellar findings reported here, but they do not establish a common causal mechanism.

**Figure 3.**
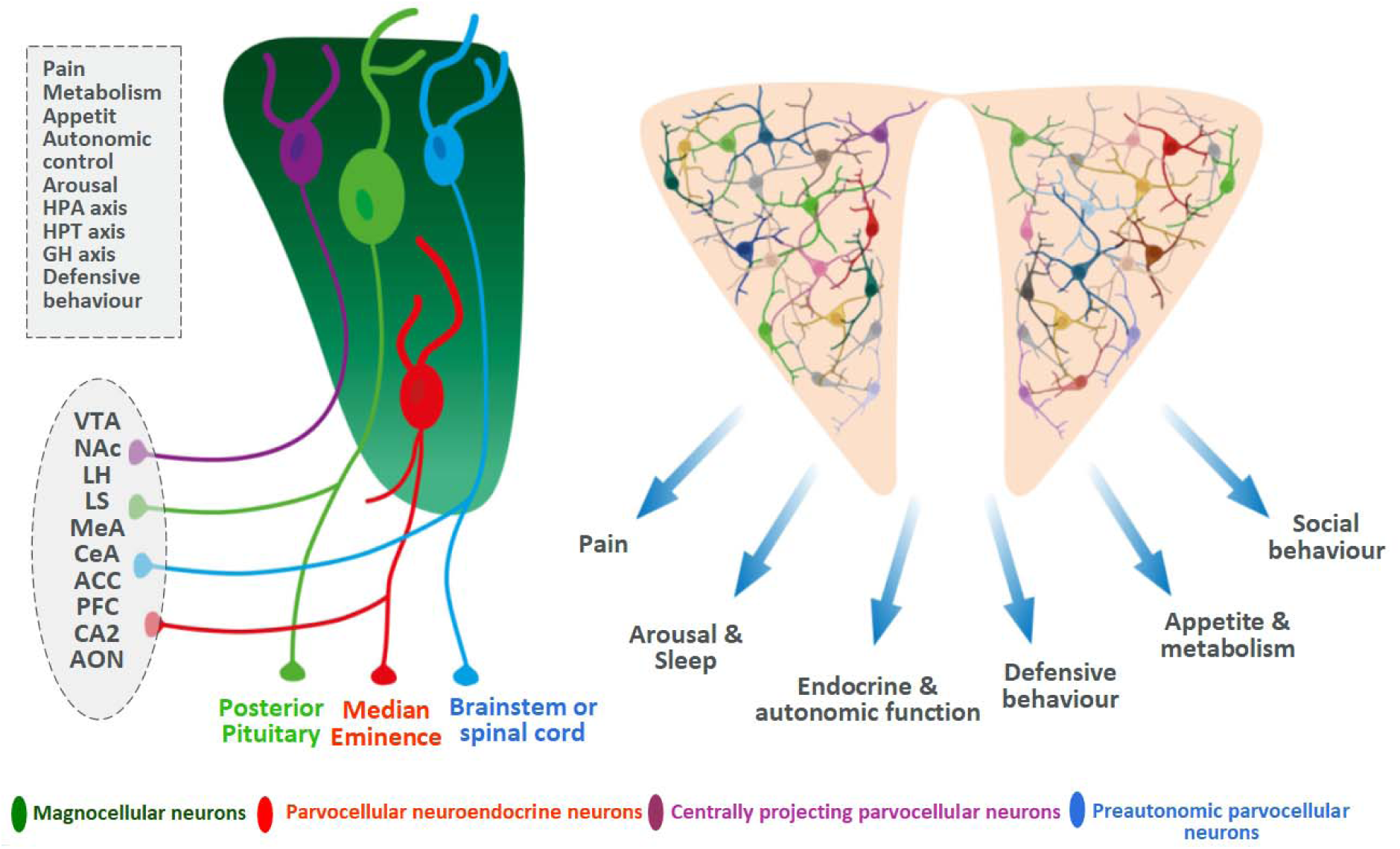
Schematic representation of superior tuberal and paraventricular hypothalamic organisation. The left panel illustrates neuronal populations and projections involved in pain, metabolism, appetite, autonomic control, arousal, and hypothalamic-pituitary regulation. The right panel summarises major functional outputs of the paraventricular region. This schematic provides anatomical context and is not a direct study result.

**Figure 4.**
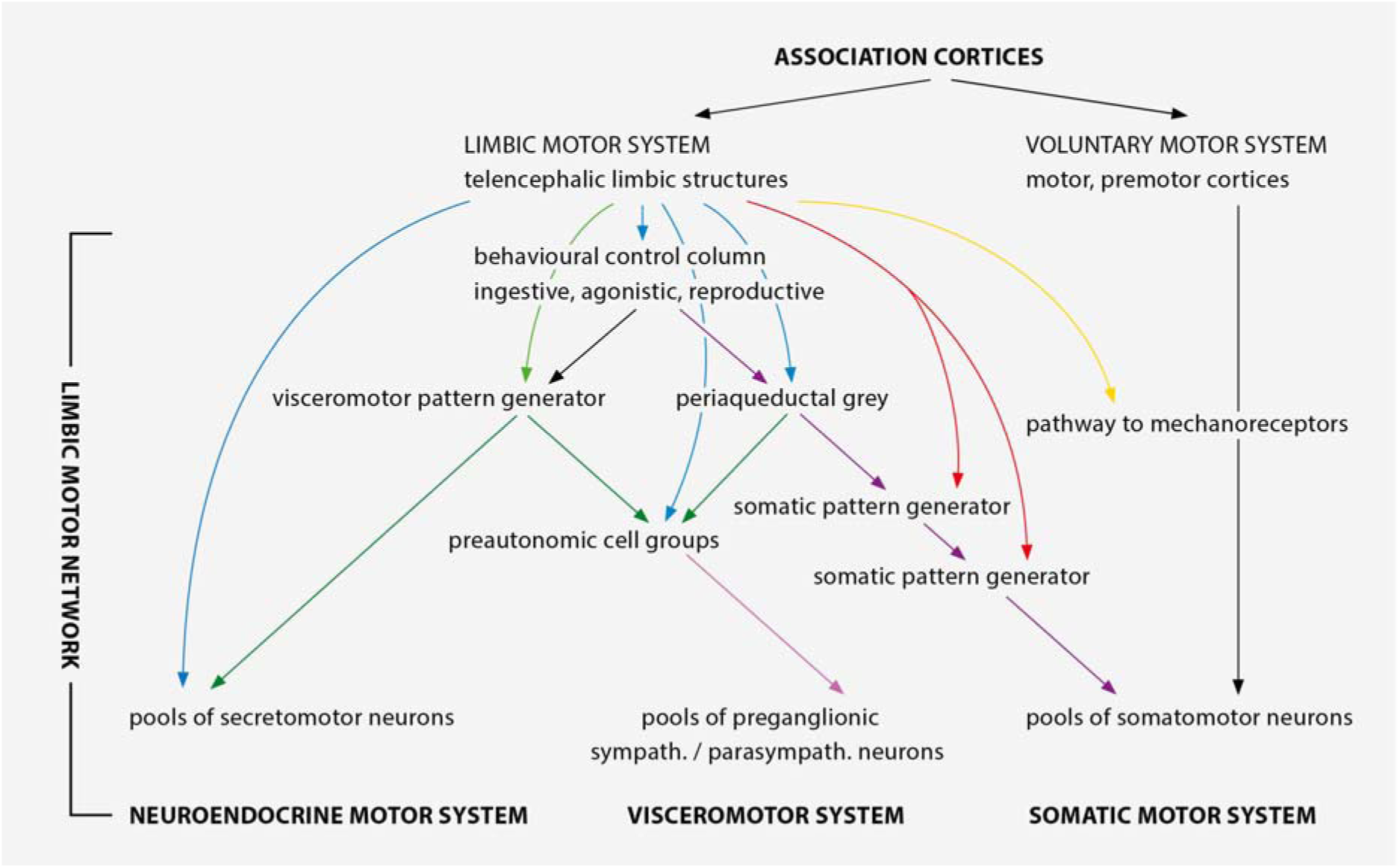
Schematic organisation of the limbic motor network and its relationships with voluntary, visceromotor, autonomic, and neuroendocrine systems. The hypothalamic visceromotor pattern-generator concept is a modification and based on the anatomical framework described by Thompson and Swanson (2003). The figure is included to illustrate the broader neuroanatomical rationale and does not represent a human Long COVID-specific pathway. With permission from the authors.

## 3. Literature context

### 3.1 Long COVID neuroimaging findings

Structural, metabolic, and diffusion imaging studies support central nervous system involvement in at least some patients with Long COVID. Longitudinal UK Biobank data demonstrated post-infection structural changes concentrated in olfactory-connected and parahippocampal regions, although the cohort was not selected for persistent symptoms (Douaud et al., 2022). FDG-PET studies in Long COVID and post-acute sequelae of COVID-19 have reported hypometabolism involving combinations of parahippocampal or mesiotemporal regions, thalamus, brainstem, frontobasal cortex, olfactory structures, and cerebellum (Guedj et al., 2021). These metabolic patterns support involvement of related limbic, diencephalic, and brainstem regions, but they do not define a directional transmission pathway.

Diffusion MRI studies have reported widespread and tract-specific white-matter differences after COVID-19. Limbic tracts have shown microstructural alterations in symptomatic post-COVID cohorts, while other studies have identified broader white-matter changes compatible with several possible tissue processes (Newcombe et al., 2023; Rau et al., 2024). Longitudinal work also indicates that diffusion measures may change over time and may partially recover in some patients, cautioning against interpreting a cross-sectional reduction in fractional anisotropy as fixed or progressive damage (Huang et al., 2023).

Recent studies further suggest that Long COVID is neurobiologically heterogeneous. Structural brain changes have been linked to cognitive outcomes in some cohorts, whereas a recent limbic TSPO-PET study did not identify uniformly increased glial activation compared with controls, despite symptom associations within the patient group (Pacheco-Jaime et al., 2025; Tuomaala et al., 2026). Resting-state work has identified altered limbic-default-mode network dynamics, again supporting network-level dysfunction rather than a single lesion model (Cahart et al., 2026).

### 3.2 Barrier, immune, and autonomic context

Blood-brain barrier and choroid plexus findings provide a biologically plausible route through which systemic immune and vascular abnormalities could influence periventricular structures. Blood-brain barrier disruption has been associated with cognitive impairment in Long COVID, and choroid plexus enlargement has been linked to cognitive and brain changes (Diez-Cirarda et al., 2025; Greene et al., 2024). A experimental mouse study indicates that SARS-CoV-2-related inflammatory processes can impair blood-brain and blood-CSF barrier function (Qiao et al., 2024)). These findings justify investigation of periventricular hypothalamic anatomy but do not demonstrate direct infection of the mammillary bodies or paraventricular nucleus.

Persistent complement activation and thromboinflammatory signalling have been reported in active Long COVID (Cervia-Hasler et al., 2024). Autoantibodies, including antibodies directed at G-protein-coupled receptors, have also been described in subsets of patients, although assay standardisation, specificity, and causality remain unresolved (Wallukat et al., 2021). Cardiovascular autonomic dysfunction is a major clinical phenotype and may include postural orthostatic tachycardia syndrome, inappropriate sinus tachycardia, hypotensive responses, impaired heart-rate and blood-pressure regulation, and microvascular manifestations (Fedorowski et al., 2024).

In post-infectious ME/CFS, Nakatomi et al. (2014) reported increased TSPO-PET signal in the thalamus, amygdala, hippocampus, cingulate cortex, and midbrain, although the study was small and used a first-generation tracer. Deep phenotyping has identified differences across central autonomic, motor-effort, immune, and metabolic systems (Walitt et al., 2024), and more recent work suggests altered central noradrenergic function in post-infectious fatigue states (Aregawi et al., 2026). Taken together, these findings support a multisystem neuroimmune-autonomic context for the present anatomical hypothesis.

## 4. Methods and materials

### 4.1 Study design and participants

This was an exploratory cross-sectional structural and diffusion MRI study including 88 participants with clinician-diagnosed Long COVID/post-COVID condition and 34 healthy controls (total N = 122). Thirty-three patients were classified as bedridden. Participants were assessed following persistent symptoms after confirmed or probable SARS-CoV-2 infection, with alternative diagnoses considered during clinical evaluation. Vaccination status, reinfection history, and medication use were documented. The study was approved by the Ethics Committee of the Hamburg Medical Association (2022-100867-BO-ff), and all participants provided informed consent.

Long COVID was defined clinically by persistent symptoms following SARS-CoV-2 infection that were not better explained by another condition. Eligibility and exclusion procedures considered neurological, endocrine, autoimmune, cardiovascular, psychiatric, sleep-related, and medication-related conditions that could materially affect the imaging or physiological outcomes.

### 4.2 MRI acquisition and preprocessing

High-resolution T1-weighted MRI was used for volumetric analysis, and diffusion MRI was used to derive diffusion-tensor measures and tractography. Imaging was processed using FreeSurfer 8.2, FSL, the CONN toolbox, and AutoPTX/Tracula as applicable. Fractional anisotropy served as the principal index of directional diffusion organisation.

Preprocessing included skull stripping, brain extraction, motion and eddy-current correction, tensor fitting, segmentation, surface reconstruction, and tractography. Quality control focused particularly on motion, registration, partial-volume effects, and segmentation reliability because the mammillary bodies, hypothalamic subregions, and brainstem nuclei are small structures.

#### 4.2.1 Image acquisition

Magnetic resonance imaging (MRI) data were acquired using a 3 Tesla Siemens Skyra scanner (Siemens Healthcare, Erlangen, Germany) at the University Medical Center Hamburg-Eppendorf. The imaging protocol included:

1. T1-weighted MPRAGE: A high-resolution T1-weighted magnetization-prepared rapid gradient echo (MPRAGE) sequence was acquired for volumetric analysis. Sequence parameters were: TR = X ms, TE = Y ms, TI = Z ms, flip angle = A degrees, voxel size = B x C x D mm³, matrix size = E x F x G.
2. Diffusion Tensor Imaging (DTI): DTI data were acquired using a single-shot echo-planar imaging (EPI) sequence. Sequence parameters were: TR = X ms, TE = Y ms, number of diffusion directions = N, b-value = M s/mm², voxel size = O x P x Q mm³, matrix size = R x S x T.
3. The numerical values of the sequence parameters for ID 6 in t1_mprage_cor_ND (256 DICOM images) and ID 10 in ep2D_diff_tract_DTI_ORIG (65 images out of 1239). Sequence parameters of the synthetic FLASH images used by FreeSurfer (mri synthesis, mri_ms_fitparms) are TR, TE, and Flip Angle. Dimensions: 256 x 256 x 256 Voxel sizes: 1.000000, 1.000000, 1.000000; Type: UCHAR (0); FOV: 256.000; DFO: 1; Xstart: -128.0, Xend: 128.0; Ystart: -128.0, Yend: 128.0; Zstart: -128.0, Zend: 128.0; TR: 2000.00 msec, TE: 2.98 msec, TI: 900.00 msec, Flip Angle: 9.00 degrees; Nframes: 1.

##### Volumetric Analysis

Volumetric analysis of the fornix and hypothalamus structures was performed using FreeSurfer version 8.2 (http://surfer.nmr.mgh.harvard.edu/). This open-source software package provides automated procedures for segmentation, surface reconstruction, and volume estimation of various brain regions (Iglesias et al., 2015). The T1-weighted MPRAGE images were processed through the standard FreeSurfer pipeline, which includes:

1. Skull stripping: Removal of non-brain tissue.
2. Segmentation: Identification and labeling of different brain structures based on their anatomical properties.
3. Surface Reconstruction: Creation of 3D models of the cortical and subcortical surfaces.
4. Volume Estimation: Calculation of the volume of each segmented brain region.

Diffusion Tensor Imaging (DTI) Analysis

DTI data were preprocessed using the FMRIB Software Library (FSL, version 6.0.7.19). The preprocessing steps included:

1. Eddy Current Correction: Correction for distortions caused by eddy currents and subject motion.
2. Brain Extraction: Removal of non-brain tissue.
3. Diffusion Tensor Calculation: Estimation of the diffusion tensor at each voxel.

From the diffusion tensor, fractional anisotropy (FA) and mean diffusivity (AD) maps were calculated. FA, a measure of white matter integrity, reflects axonal fiber density, axonal diameter, and myelination. The FA_Avg fractional anisotropy average testing was performed using FreeSurfer Tracula, average conductivity in the vector areas 45 to 85 shown in boxplots. The average FA value was extracted from the fornix, as this region demonstrated significant volume and conductivity reductions in the analysis.

### 4.3 Regions of interest and operational definitions

Primary regions of interest were the left and right mammillary bodies, the superior tuberal/periventricular hypothalamic region, and the fornix. Outcomes included regional volume, fornix fractional anisotropy and axial diffusivity, tract coherence, and the visually identified narrowing or loss of a visible internal passage at the superior tuberal-mammillary interface.

**Figure 5.**
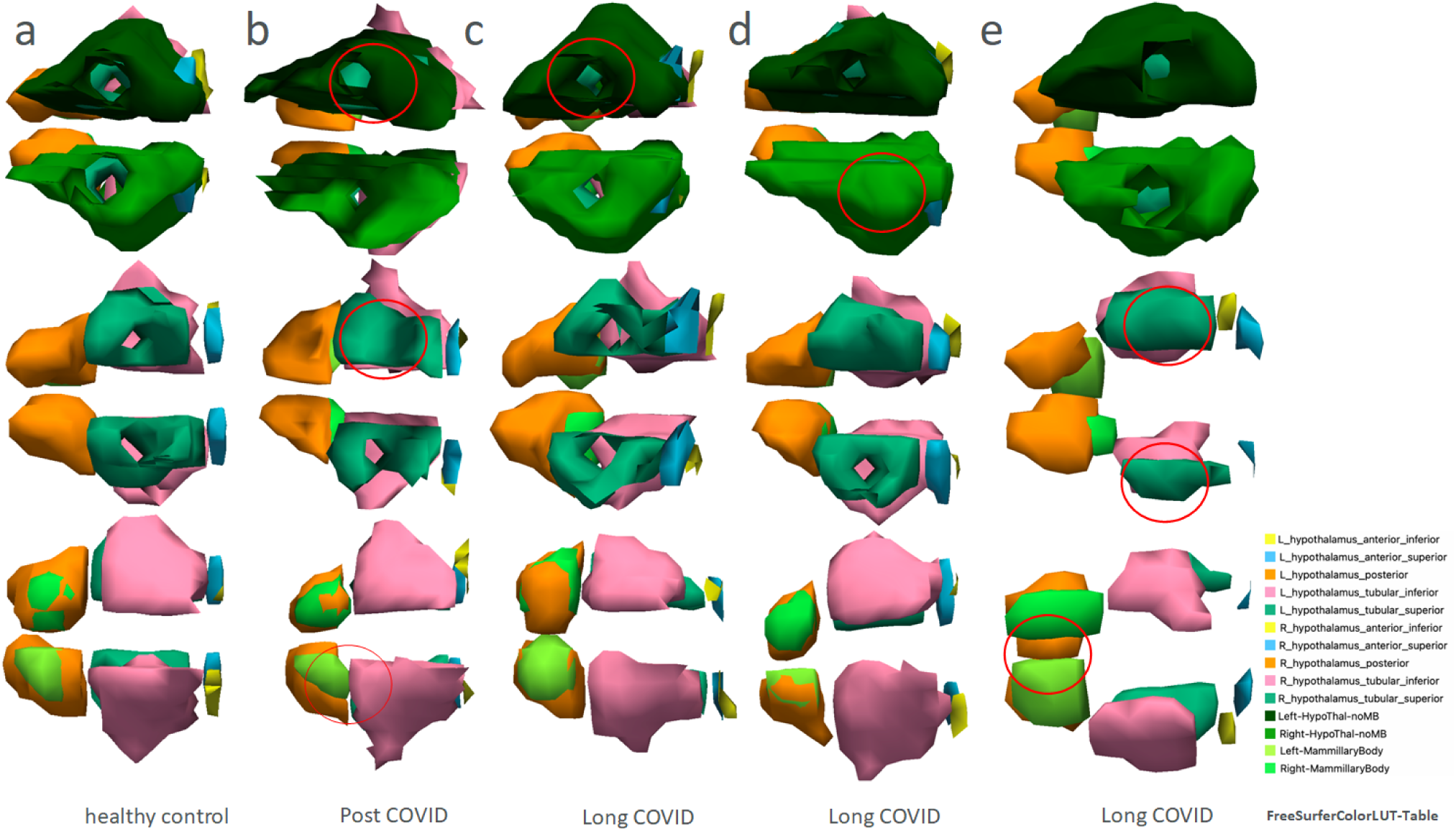
Three-dimensional FreeView isosurface examples of hypothalamic segmentation in a healthy control (a) and participants with post-COVID/Long COVID (b-e). The mammillary body is shown in bright green, the inferior tuberal region in pink, the superior tuberal region in green, and the HypoThal-noMB region in dark green. Red circles indicate regions in which the visible separation between adjacent segmented structures is reduced or absent. These images illustrate morphological segmentation differences without establishing inflammation, compression, or tissue destruction.

Secondary analyses examined fornix volume and diffusion measures across segments approaching the superior and inferior tuberal regions and mammillary bodies. These analyses were treated as exploratory and interpreted separately from the primary gate hypothesis.

**Figure 6.**
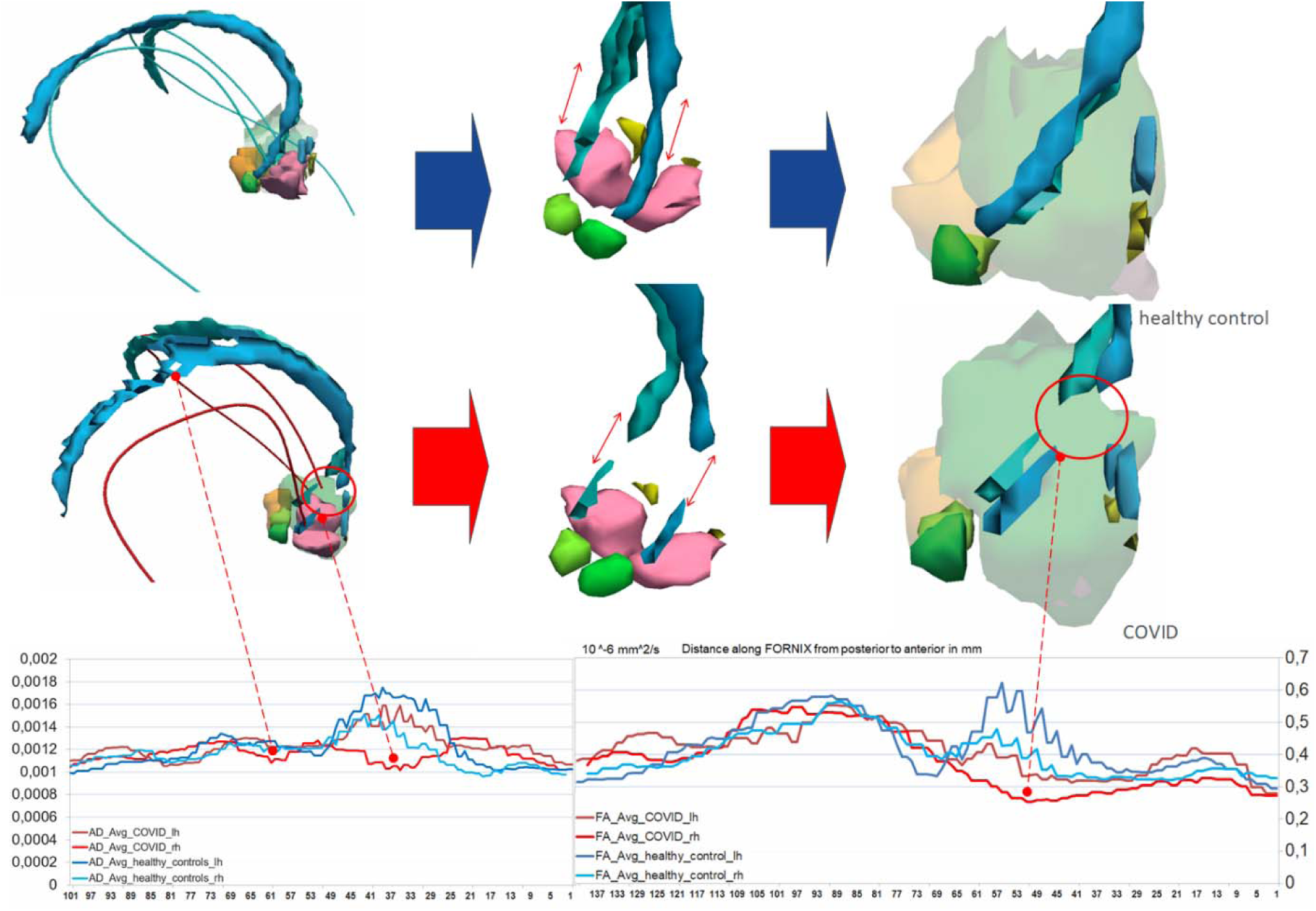
Three-dimensional fornix and hypothalamic reconstructions with along-tract fractional anisotropy and axial-diffusivity profiles. The upper row illustrates a healthy-control reconstruction and the lower row a Long COVID reconstruction. The Long COVID example shows attenuation or discontinuity of the reconstructed fornix segment approaching the superior tuberal-mammillary region. Because tractography is sensitive to acquisition, partial-volume effects, crossing fibres, and tracking thresholds, the image is interpreted as altered tract reconstruction rather than proof of axonal destruction or mechanical disconnection.

**Figure 6b.**
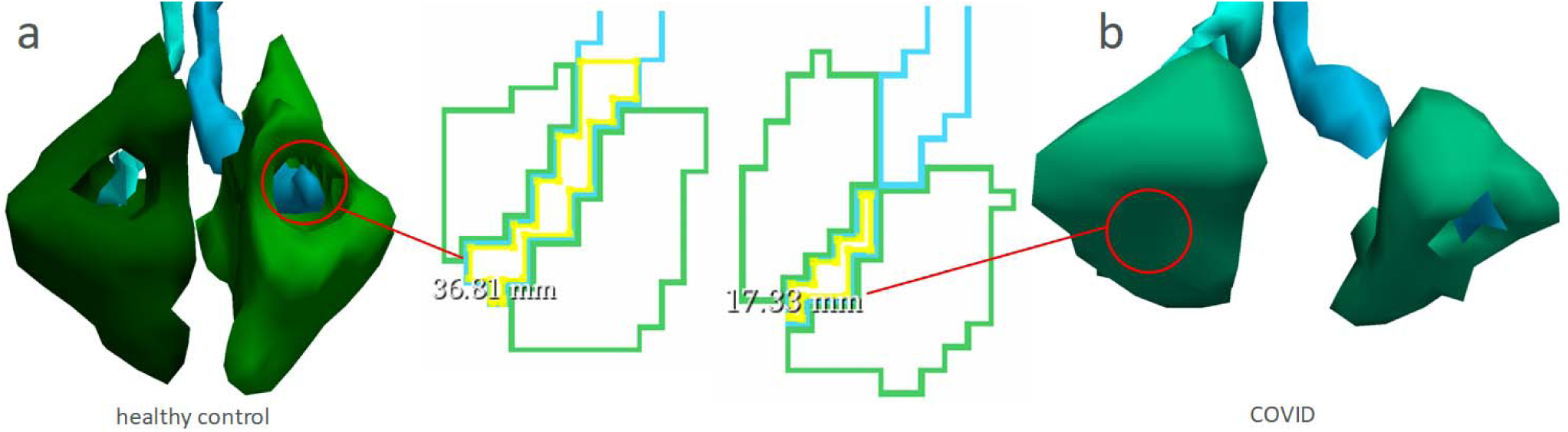
Inferior views of the bilateral fornix as it approaches the hypothalamic plane. Panel a shows a healthy-control example and panel b a Long COVID example, including serial images from 2023 to 2026 in one participant. The Long COVID example shows reduced visibility of an internal passage in the tuberal region.

**Figure 6c.**
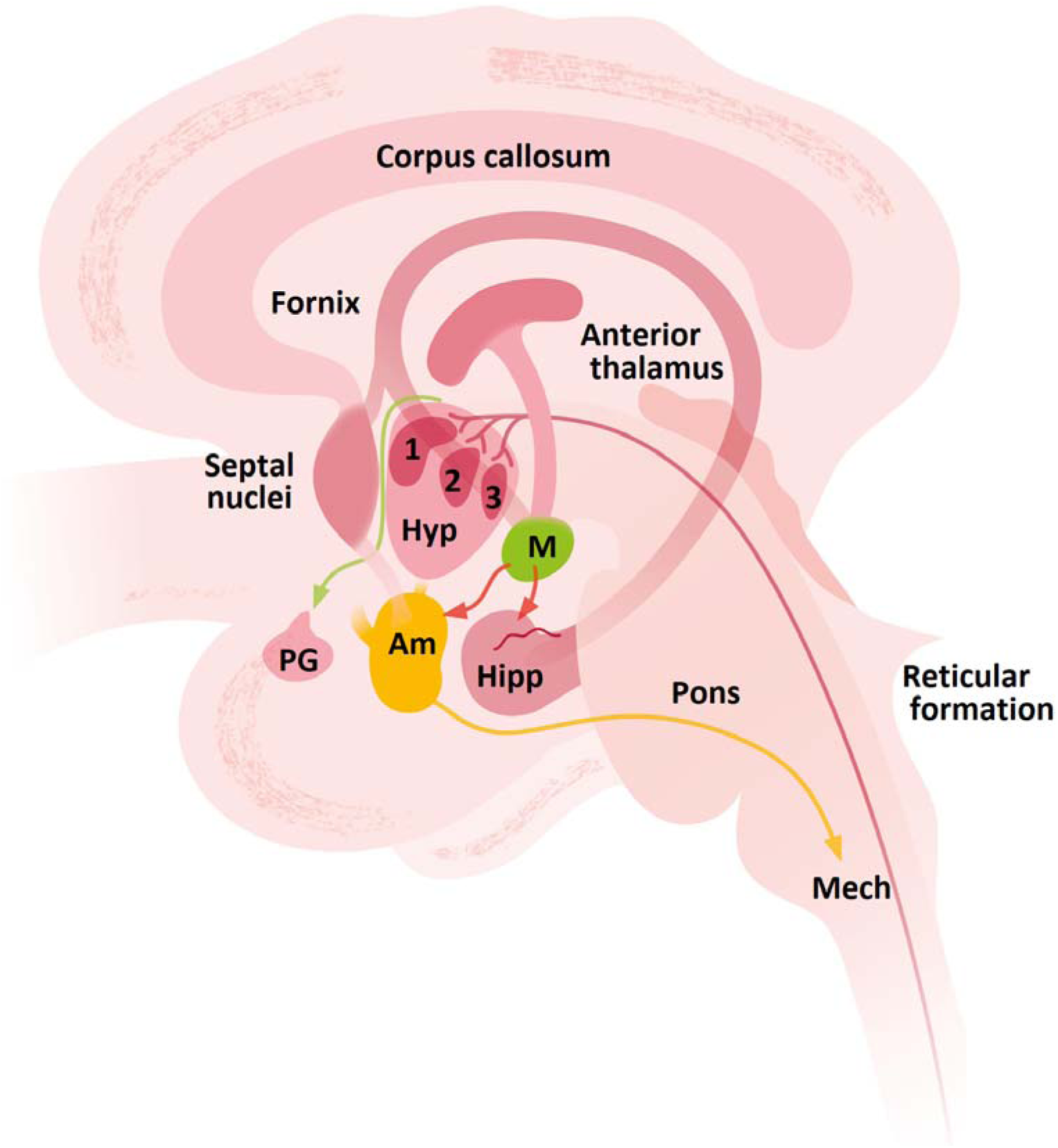
Schematic medial view of limbic and hypothalamic connections. The diagram illustrates hippocampal-forniceal projections to the mammillary body and anterior thalamus, together with hypothalamic-reticular-spinal sympathetic pathways. It is included as anatomical context and does not depict a newly established Long COVID circuit.

### 4.4 Clinical measures

Clinical and physiological assessments were performed by the same clinical research team using a standardised protocol. Long COVID and ME/CFS-related symptoms were evaluated through a semi-structured clinical interview based on the Canadian Consensus Criteria, including post-exertional malaise, fatigue, sleep disturbance, pain, cognitive symptoms, autonomic dysfunction, neuroendocrine symptoms, and immune-related symptoms. Autonomic and neuromuscular function were assessed during a 20-minute supine resting period, followed by a controlled exertion phase and recovery. Measurements included electrocardiography for heart-rate variability analysis, oxygen saturation, resting tremor or microvibration, and surface electromyography of the thigh musculature during exertion. Heart-rate variability was analysed using the root mean square of successive differences (RMSSD). Electrocardiographic signals were recorded at 1,000 Hz and processed using Nerve-Express and Kubios HRV Premium; R-peak detection was visually checked and corrected for artefacts. Participants were instructed to avoid alcohol, nicotine, and caffeine on the day before and the day of testing. Internal vibrations and tremor-like symptoms were classified according to whether they were patient-reported, clinically observed, or captured physiologically.

### 4.5 Statistical analysis

Statistical analyses were performed using SPSS version 18 and R. Group comparisons of regional volumes and diffusion measures used independent-samples tests or one-way analysis of variance, with Welch procedures where variance assumptions were not met and Bonferroni-adjusted post-hoc comparisons for the three mammillary-body groups. Effect estimates included partial eta-squared and Hedges’ g. Pearson correlations were used for exploratory associations between imaging measures and clinical variables. Statistical significance was set at p < .05. Because multiple small regions and diffusion measures were examined, the findings were interpreted as exploratory.

**Figure 7.**
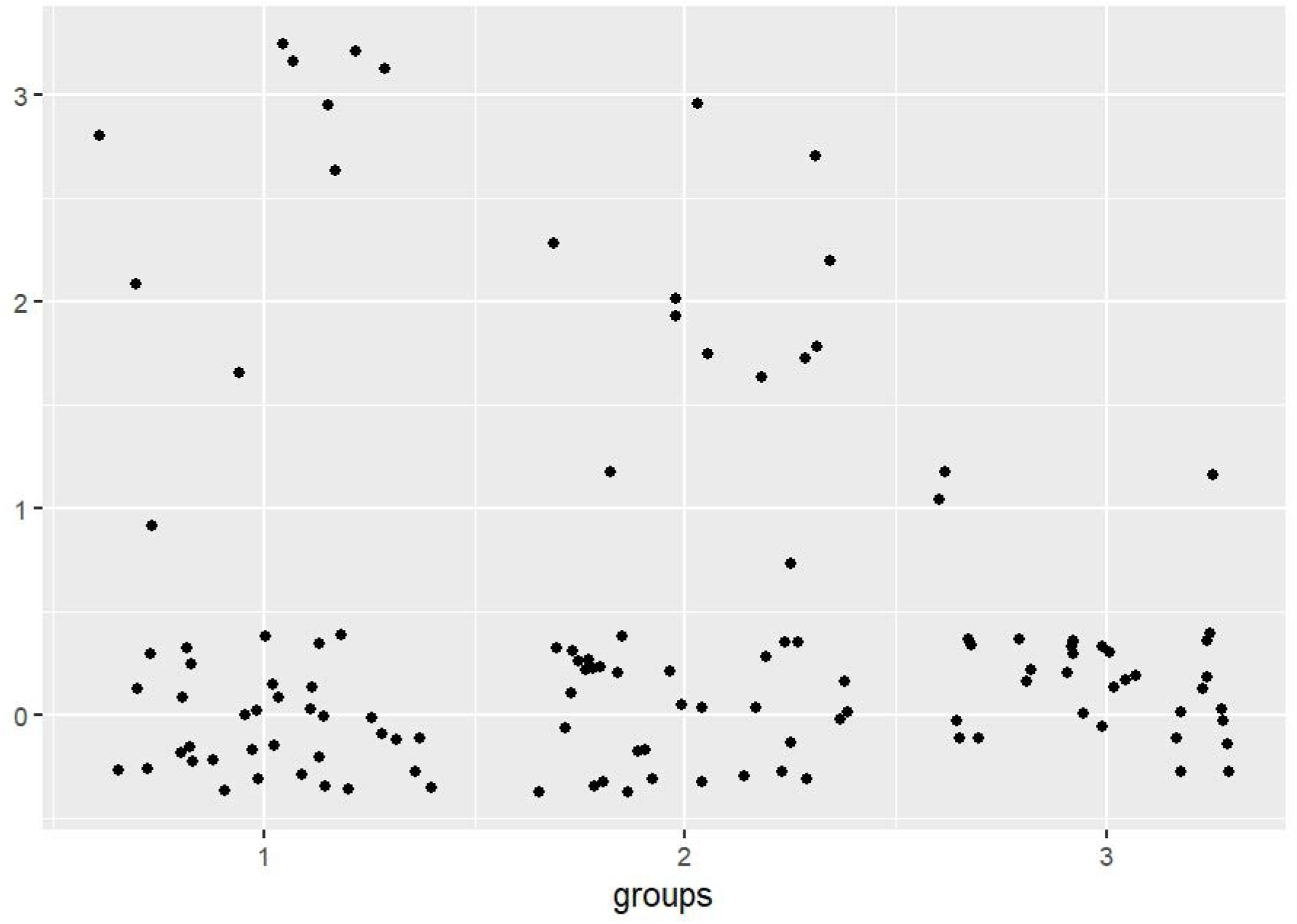
Distribution of mammillary-body and superior tuberal morphology across the study sample. The plotted points represent all 122 participants. Group 1 comprised participants with reduced mammillary-body volume, Group 2 participants with enlarged mammillary-body volume, and Group 3 participants with values within the healthy-control range. Segmentation-defined narrowing of the tuberal internal passage was observed in 26 patients and three controls.

**Figure 8.**
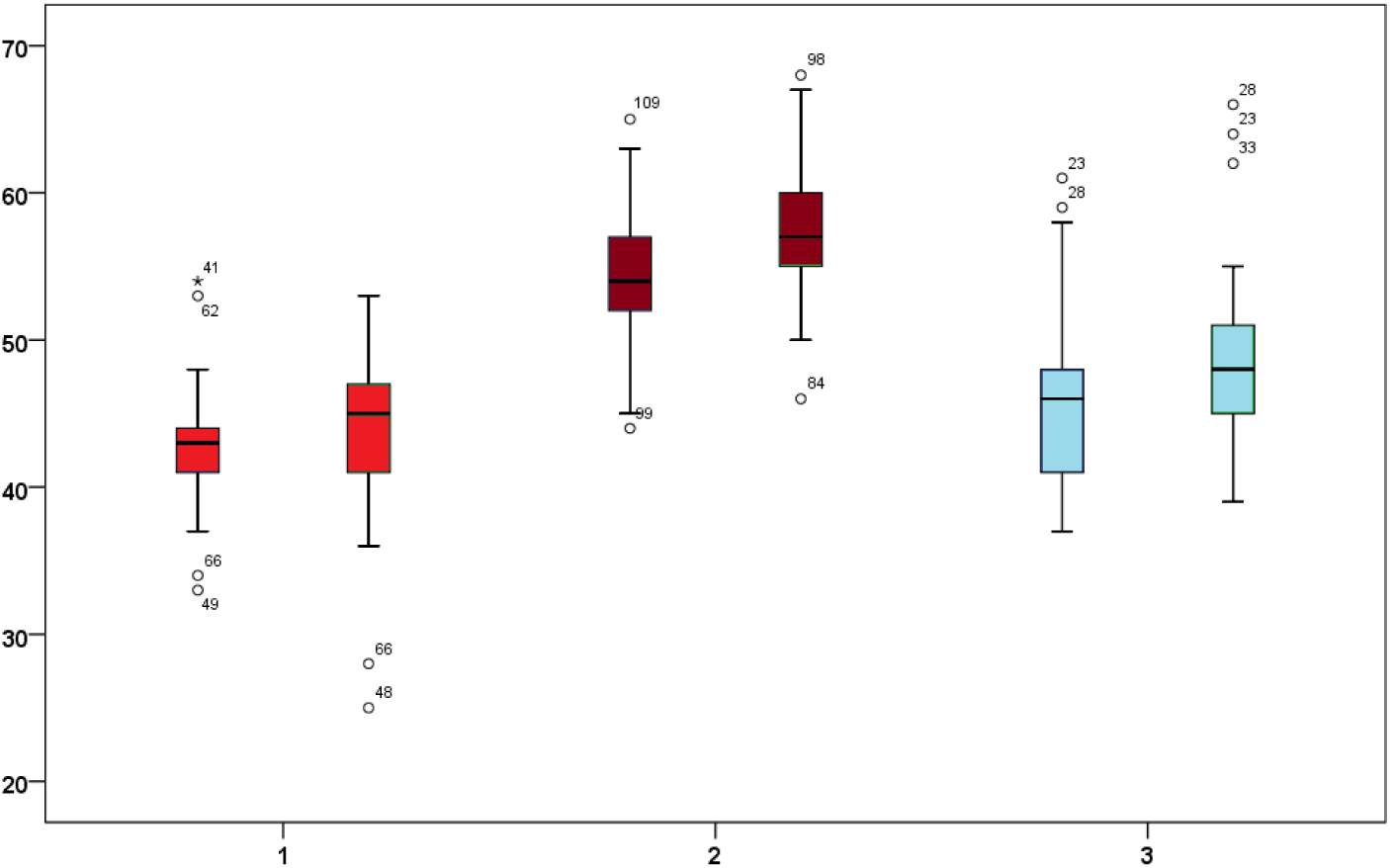
Box plots of left and right mammillary-body volumes across three phenotypic groups: reduced volume (Group 1), enlarged volume (Group 2), and control-range volume (Group 3). The distributions show separation between the three groups.

One-way analysis of variance with Bonferroni-adjusted post-hoc comparisons identified differences across the three mammillary-body groups. Partial eta-squared was .554 for the left mammillary body and .559 for the right mammillary body, indicating large between-group effects in this exploratory analysis.

**Figure 8b.**
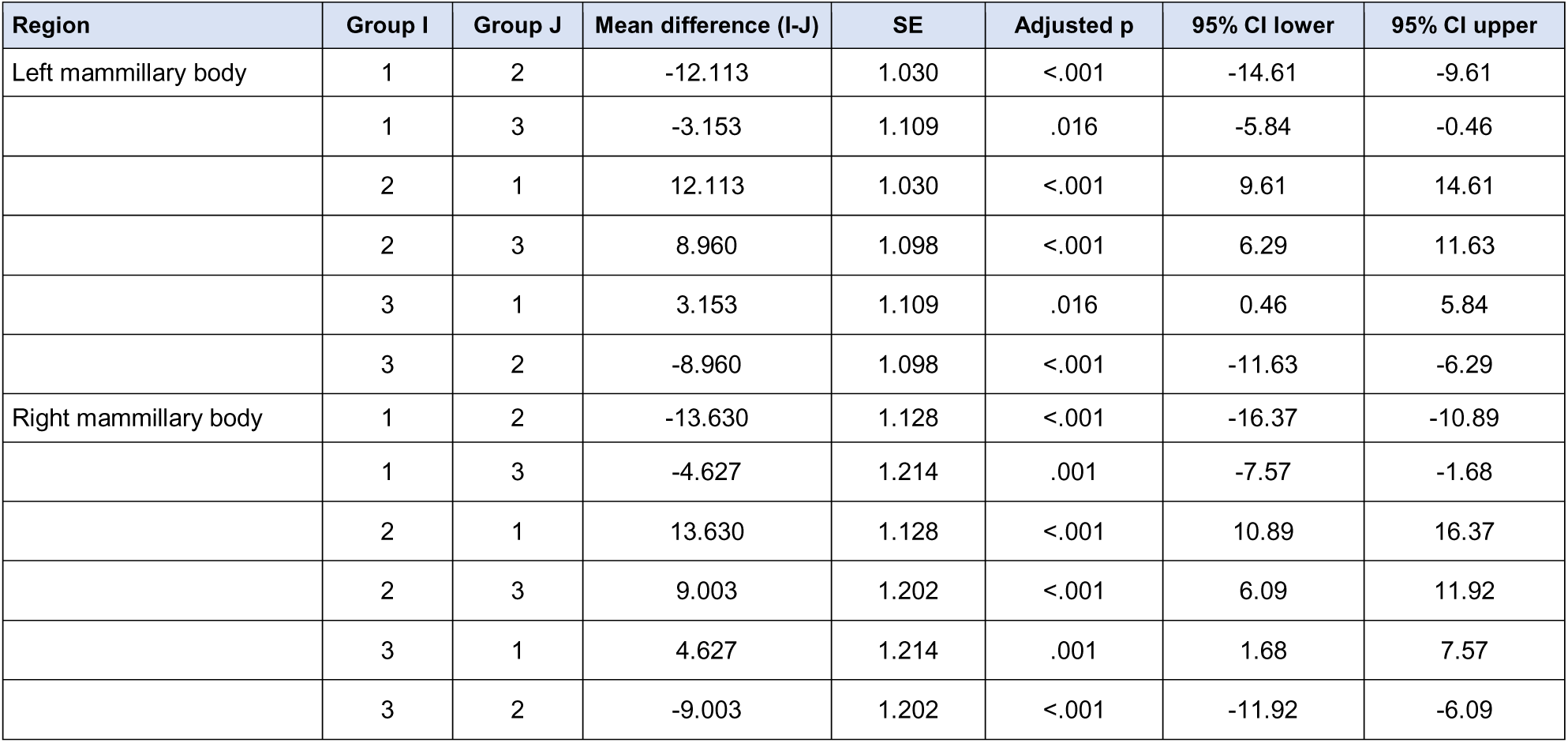
Bonferroni-adjusted pairwise comparisons of left and right mammillary-body volumes across the three phenotypic groups. The table displays mean differences, standard errors, adjusted significance values, and 95% confidence intervals.

**Figure 9.**
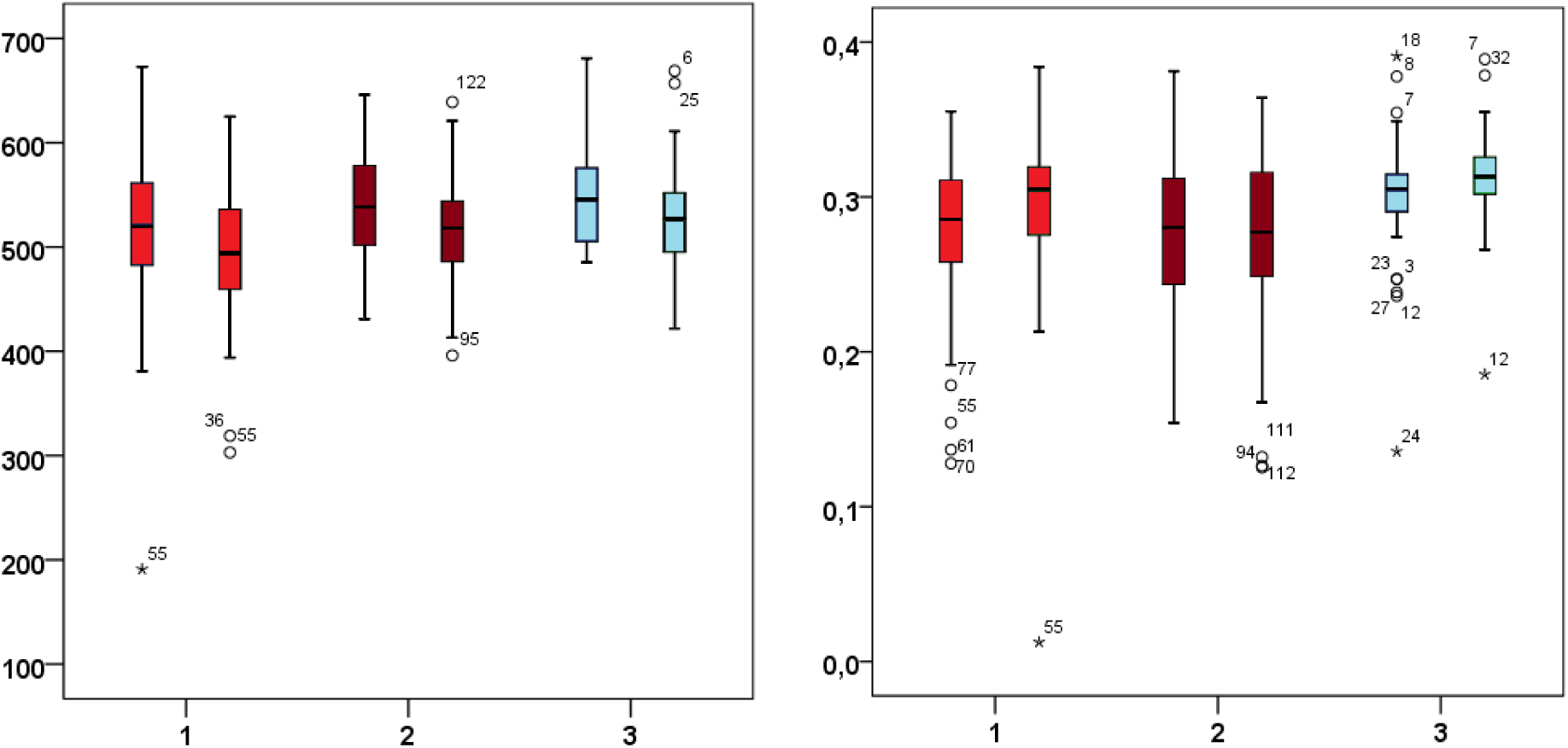
Fornix volume and fractional-anisotropy distributions across the two Long COVID subgroups and healthy controls. The eft panel shows regional fornix volume; the right panel shows average fractional anisotropy across the specified anterior fornix segment. The visualisation is descriptive and does not depict a demonstrated route of viral spread.

**Table 2.**
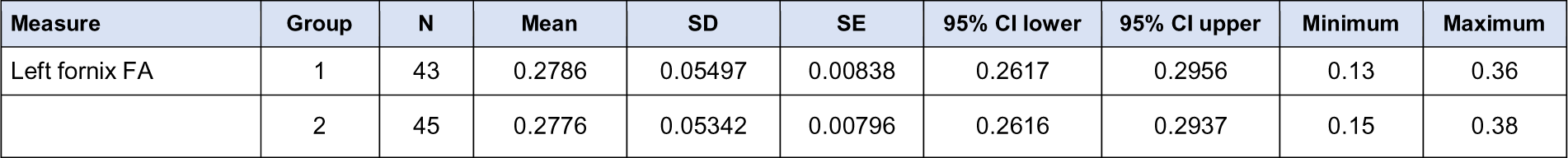
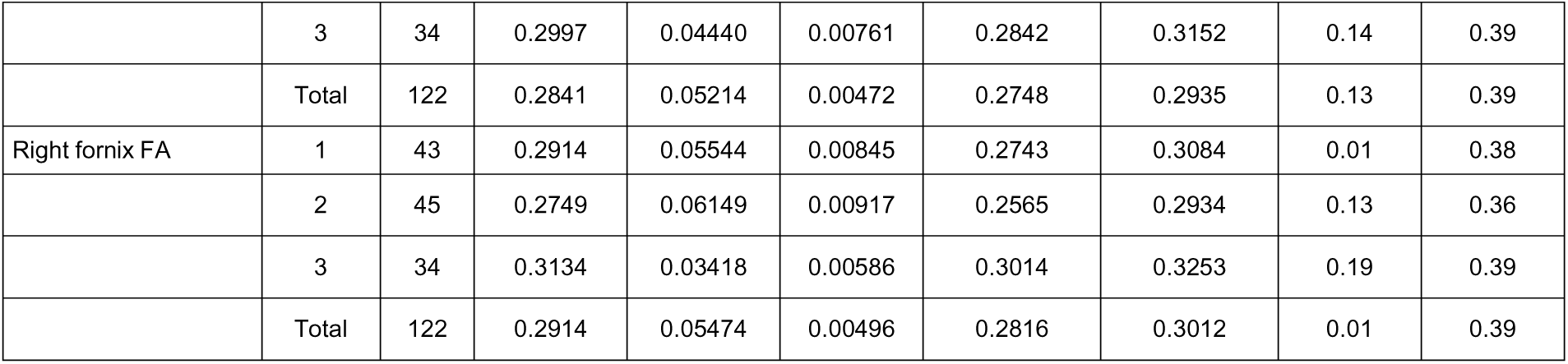
Descriptive statistics for left and right fornix fractional anisotropy across the three groups. The Welch test identified a right-sided group difference (p = .002). Post-hoc comparison indicated higher right fornix fractional anisotropy in healthy controls than in the enlarged-volume Long COVID subgroup.

**Figure 10b.**
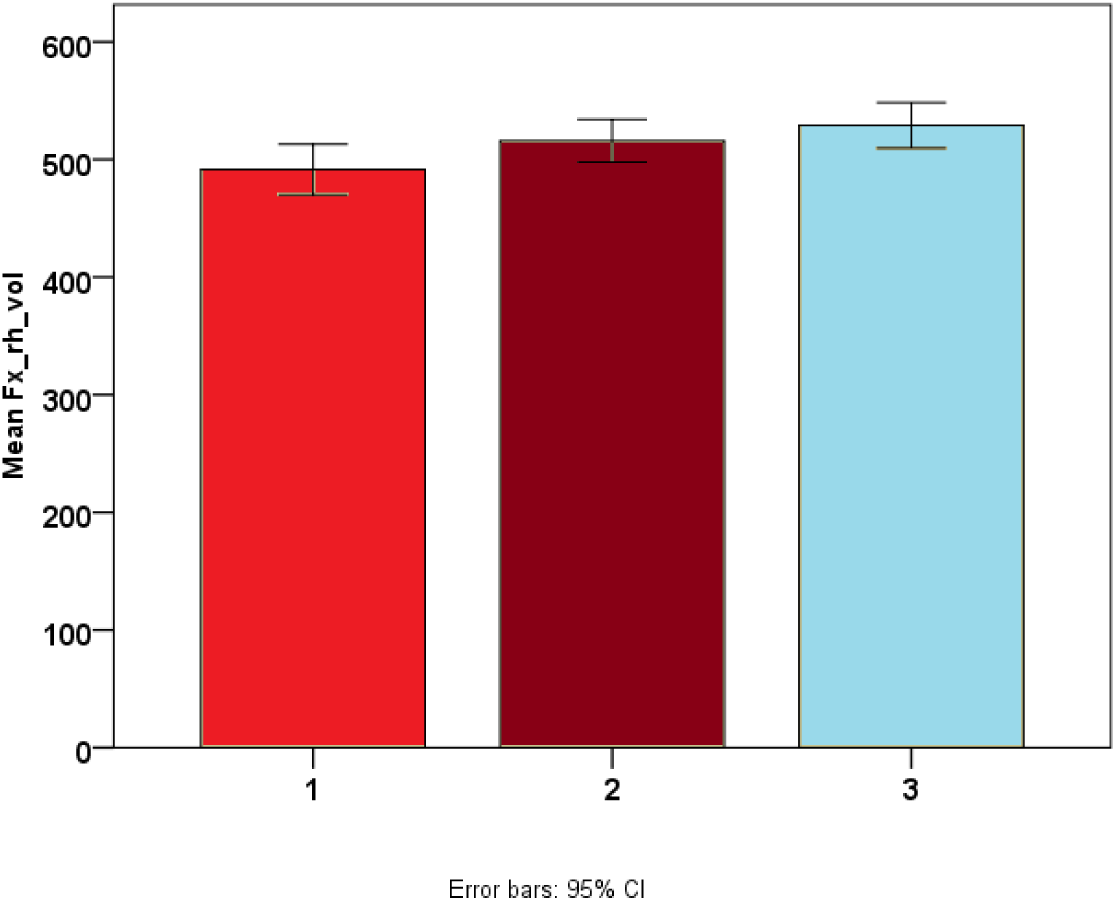
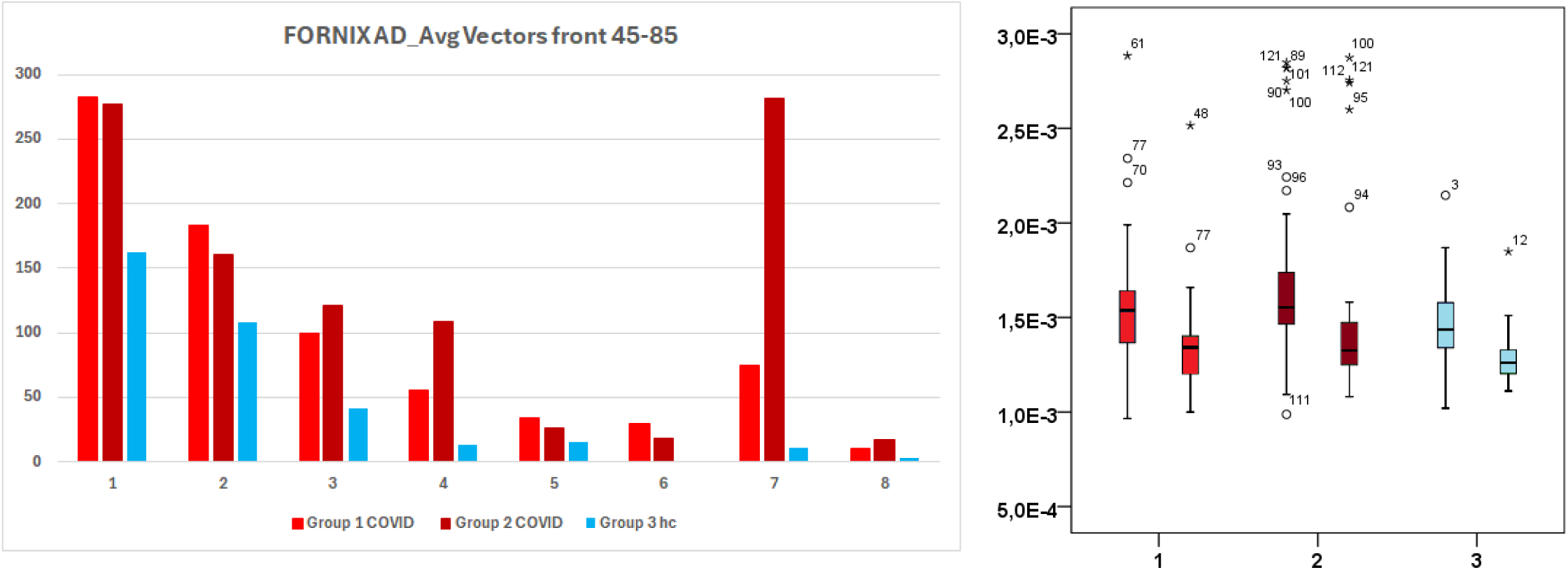
Mean right fornix volume with 95% confidence intervals across the three groups. Error bars show uncertainty around each group mean and should not be interpreted as measures of individual variability.

**Table 3.**
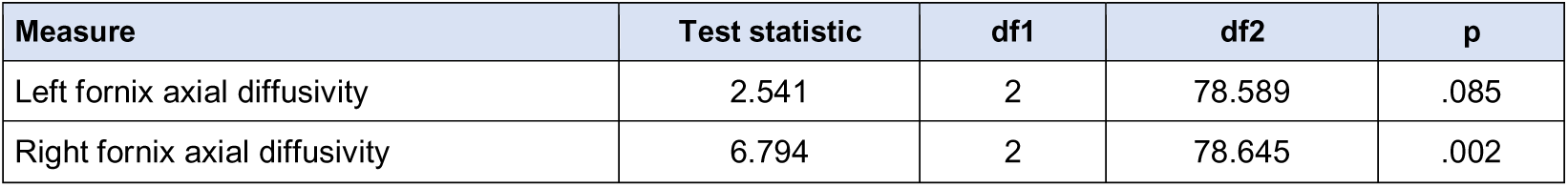
Welch tests for group differences in left and right fornix axial diffusivity.

**Table 4.**
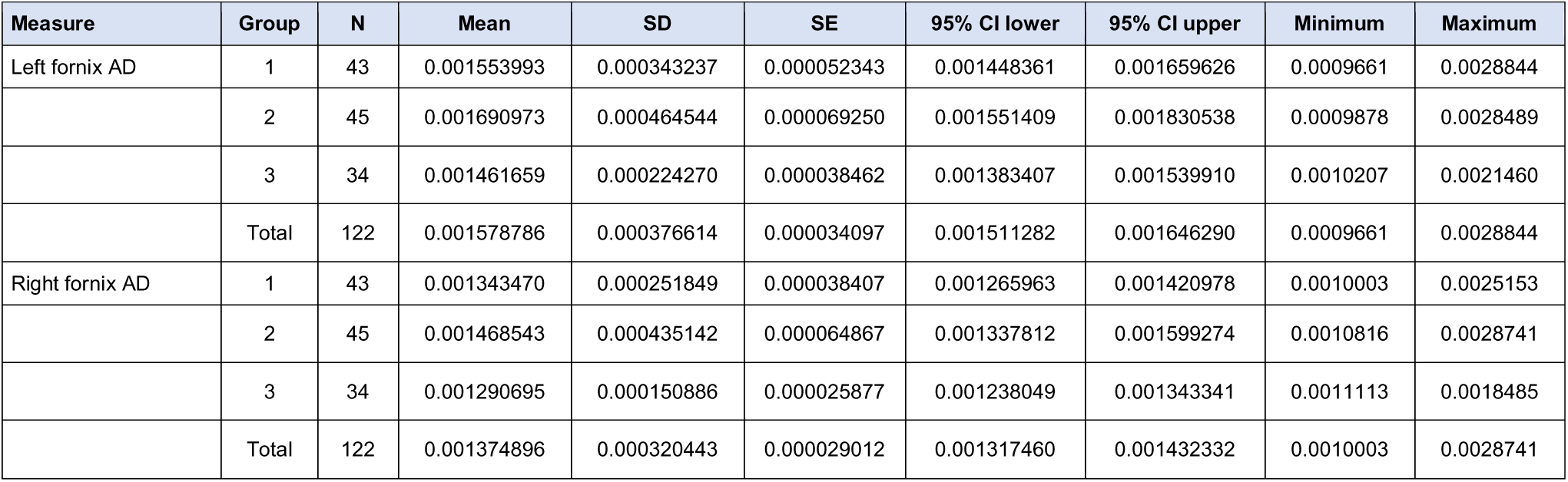
Descriptive statistics for left and right fornix axial diffusivity across the three groups.

| Measure | Group | N | Mean | SD | SE | 95% CI lower | 95% CI upper | Minimum | Maximum |
| --- | --- | --- | --- | --- | --- | --- | --- | --- | --- |
| Left fornix AD | 1 | 43 | 0.001553993 | 0.000343237 | 0.000052343 | 0.001448361 | 0.001659626 | 0.0009661 | 0.0028844 |
|  | 2 | 45 | 0.001690973 | 0.000464544 | 0.000069250 | 0.001551409 | 0.001830538 | 0.0009878 | 0.0028489 |
|  | 3 | 34 | 0.001461659 | 0.000224270 | 0.000038462 | 0.001383407 | 0.001539910 | 0.0010207 | 0.0021460 |
|  | Total | 122 | 0.001578786 | 0.000376614 | 0.000034097 | 0.001511282 | 0.001646290 | 0.0009661 | 0.0028844 |
| Right fornix AD | 1 | 43 | 0.001343470 | 0.000251849 | 0.000038407 | 0.001265963 | 0.001420978 | 0.0010003 | 0.0025153 |
|  | 2 | 45 | 0.001468543 | 0.000435142 | 0.000064867 | 0.001337812 | 0.001599274 | 0.0010816 | 0.0028741 |
|  | 3 | 34 | 0.001290695 | 0.000150886 | 0.000025877 | 0.001238049 | 0.001343341 | 0.0011113 | 0.0018485 |
|  | Total | 122 | 0.001374896 | 0.000320443 | 0.000029012 | 0.001317460 | 0.001432332 | 0.0010003 | 0.0028741 |

To visualize structural changes, 3D reconstructions of the brainstem regions of interest were generated using FreeSurfer’s isosurface reconstruction tool and isosurface 3D view, and Horos data transformation. This approach allowed for the visualization and validation of the observed structural and functional abnormalities.

## 5. Results

### 5.1 Mammillary-body and superior tuberal phenotypes

Three mammillary-body volume phenotypes were identified: reduced volume, enlarged volume, and control-range volume. Differences were observed for both left and right mammillary-body volumes, with large partial eta-squared estimates (left ηp² = .554; right ηp² = .559). Three-dimensional segmentation also showed morphological differences in the superior tuberal/periventricular region, including narrowing or loss of a visible internal passage adjacent to the mammillary body-fornix interface (Fig. 5 and 6). This is reported as a segmentation-defined morphological finding and does not establish direct forniceal compression, obstruction, inflammation, or tissue destruction.

Three-dimensional segmentation showed morphological differences in the superior tuberal/periventricular region, including narrowing or loss of a visible internal passage adjacent to the mammillary body-fornix interface. This finding is described as a morphological abnormality. It may represent a structural bottleneck, but the imaging data do not establish direct forniceal compression, obstruction, or tissue destruction.

### 5.2 Fornix diffusion findings

The fornix showed altered diffusion measures and reduced tract coherence in the hypothesised gate region (Fig. 6). A right-sided group difference was reported using Welch’s test (p = .002), with healthy controls showing higher right fornix fractional anisotropy than one Long COVID subgroup (Table 3). . The tractography images suggested an attenuated or discontinuous segment approaching the mammillary bodies.

These observations are consistent with altered forniceal microstructure. Fractional anisotropy and tractography are influenced by fibre coherence, crossing fibres, partial-volume effects, edema, myelination, acquisition quality, and tracking parameters; they do not directly demonstrate axonal destruction.

### 5.3 Brainstem-cerebellar findings

Long COVID participants showed lower superior cerebellar peduncle volume than controls (219.74 mm³ vs. 347.03 mm³, p < .001; Hedges’ g = 3.31). Middle cerebellar peduncle fractional anisotropy was also lower (0.39 vs. 0.45, p < .001; Hedges’ g = 1.77). Lower dorsal raphe and midbrain reticular formation volumes were additionally observed. These findings identify a second cluster involving brainstem arousal and cerebellar-motor pathways. The large effects estimates require cautious interpretation because these small structures are technically challenging to segment and are sensitive to registration and motion artefacts.

These findings identify a second cluster involving brainstem arousal and cerebellar-motor pathways. The large effect estimates require careful quality control and independent replication because small brainstem nuclei and cerebellar peduncles are technically challenging to segment and are sensitive to registration and motion artefacts.

### 5.4 Clinical associations

Exploratory associations were observed between regional volume or diffusion abnormalities and motor deficits, proprioceptive dysfunction, autonomic dysregulation, fatigue, internal vibrations, and tremor-like symptoms.

The study sample comprised 122 participants, who were classified into three phenotypic groups according to mammillary body volume: reduced volume (Group 1), enlarged volume (Group 2), and volumes within the healthy control range (Group 3). Segmentation-defined narrowing of the tuberal internal passage was identified in 26 patients and three healthy controls.

Box plots of left and right mammillary body volumes demonstrated clear separation between the three phenotypic groups. Bonferroni-adjusted pairwise comparisons confirmed significant differences in mammillary body volumes across groups, supporting the phenotypic classification.

For fornix microstructure, Welch’s ANOVA revealed a significant group effect for right fornix fractional anisotropy (p = 0.002). Post hoc analysis showed higher right fornix fractional anisotropy in healthy controls than in the Long COVID subgroup with enlarged mammillary body volume. Group differences in right fornix volume are presented as estimated means with 95% confidence intervals; the error bars represent the uncertainty of the estimated group means rather than individual variability.

These associations should be interpreted considering the measurement method used for each symptom and the absence of independent replication.

## 6. Discussion

### 6.1 Principal findings

This exploratory study identified a pattern of structural and diffusion abnormalities centred on the mammillary body-fornix-superior tuberal hypothalamic interface, with additional abnormalities in brainstem and cerebellar pathways. The most important finding is not a single isolated region, but the clustering of abnormalities across systems involved in hippocampal-diencephalic communication, hypothalamic homeostasis, brainstem arousal and autonomic regulation, and cerebellar-motor coordination.

The mammillary body findings were heterogeneous, with reduced, enlarged, and control-range phenotypes. This pattern argues against a single uniform lesion and is more compatible with different biological stages or subgroups, measurement variability, or distinct combinations of inflammatory, vascular, metabolic, and neurodegenerative processes. Longitudinal imaging is required to determine whether enlarged and reduced volumes represent different trajectories, transient states, or unrelated phenotypes.

### 6.2 The mammillary body-fornix gate hypothesis

The gate model proposes that the small periventricular interface between forniceal fibres, mammillary bodies, and adjacent superior tuberal hypothalamus may be vulnerable to processes that alter local morphology or diffusion organisation. In a subgroup of patients, disruption at this interface could affect communication between hippocampal-contextual systems and hypothalamic homeostatic output. Brainstem-cerebellar abnormalities may then contribute to impaired arousal regulation, postural control, autonomic adaptation, tremor-like symptoms, and motor fatigability.

This interpretation is anatomically plausible but remains inferential. The study does not show that SARS-CoV-2 or spike protein entered the brain through the anterior commissure, nor does it demonstrate direct viral infection of the hypothalamus, mammillary bodies, or fornix. The anterior commissure should therefore be treated as an observed commissural tract of interest, not as an established viral entry route. Likewise, the hypothesised pathway should be described as a set of interconnected regions rather than a formally defined single circuit.

### 6.3 Relationship to prior Long COVID and ME/CFS research

The present findings are broadly consistent with prior reports of limbic and mesiotemporal white-matter changes, thalamic and brainstem hypometabolism, brainstem MRI abnormalities, and altered subcortical connectivity after COVID-19 (Guedj et al., 2021; Rau et al., 2024; Rua et al., 2024). They also align with ME/CFS studies implicating thalamic, hippocampal, amygdalar, midbrain, and intra-brainstem systems (Barnden et al., 2019; Nakatomi et al., 2014; Shan et al., 2023). However, no prior human study has established the exact mammillary body-fornix gate model proposed here.

The literature also cautions against a simple persistent-neuroinflammation narrative. Some studies show inflammatory or barrier-associated findings, whereas others do not identify uniformly increased glial activation across Long COVID cohorts (Greene et al., 2024; Tuomaala et al., 2026). The current imaging abnormalities may therefore arise from different or interacting processes, including neuroimmune activation, endothelial dysfunction, altered perfusion, edema, demyelination, axonal change, or compensatory plasticity. The present dataset cannot distinguish among these mechanisms.

### 6.4 Clinical interpretation

Clinically, the findings support mechanism-aligned phenotyping rather than generic attribution of severe fatigue or internal vibrations to psychological causes. Patients with PEM, orthostatic intolerance, sleep-wake instability, internal vibrations or tremor-like symptoms, sensory overload, and cognitive dysfunction may have overlapping autonomic, motor, vascular, immune, and central nervous system abnormalities. The imaging findings do not provide a diagnostic biomarker at present, but they identify candidate regions for prospective validation.

The study does not justify a specific treatment. It supports future trials that stratify participants by clinical and biological phenotype and assess whether autonomic stabilisation, sleep and endocrine management, pacing within post-exertional limits, or appropriately selected immune or vascular interventions are associated with objective physiological and imaging changes.

## 7. Limitations

Several limitations substantially constrain interpretation. First, this was an exploratory analysis involving multiple small regions of interest, increasing the risk of false-positive findings. Second, segmentation of the mammillary bodies, hypothalamic subregions, raphe nuclei, reticular formation, and cerebellar peduncles is technically demanding and susceptible to partial-volume effects, motion, and registration error. Third, DTI metrics and tractography are indirect and cannot establish axonal destruction or a specific histopathological process. Fourth, the cross-sectional design precludes inference about temporal sequence or causality. Fifth, internal vibrations and tremor-like or shivering-like episodes were not uniformly characterised using objective physiological recordings. Sixth, immune, autoantibody, viral-persistence, perfusion, and barrier mechanisms were not directly demonstrated by the imaging data. Finally, the cohort was enriched for severe muscular impairment and bedridden participants, which may limit generalisability to the broader Long COVID population. Fifth, the cross-sectional design precludes inference about temporal sequence or causality. Sixth, immune, autoantibody, viral-persistence, perfusion, and barrier mechanisms were not directly demonstrated by the imaging data. Finally, Long COVID is clinically heterogeneous, and findings from a cohort enriched for severe muscular impairment or bedridden patients may not generalise to broader post-COVID populations.

## 8. Conclusions

Long COVID participants in this exploratory study showed structural and diffusion abnormalities involving the mammillary body-fornix-superior tuberal hypothalamic interface and connected brainstem-cerebellar pathways. The findings highlight a potentially important convergence zone linking hippocampal-diencephalic communication, hypothalamic homeostatic regulation, brainstem arousal and autonomic control, and cerebellar-motor coordination.

The mammillary body-fornix gate is proposed as a testable neuroanatomical model for a subgroup characterised by internal vibrations or tremor-like symptoms, neuromuscular fatigue, autonomic instability, PEM, and cognitive symptoms. The present data do not establish a viral entry route, direct infection, a thermoregulatory shivering mechanism, or a single causal pathway. Replication with standardised imaging, prespecified regions, rigorous multiplicity correction, objective physiological measures, disease-control groups, and longitudinal follow-up is required to establish the model as a biomarker or treatment target for ME/CFS pts.

## Data Availability

All data produced in the present study are available upon reasonable request to the authors
All data produced in the present work are contained in the manuscript
All data produced are available online

## Author contributions

The Authors contributed to the conception, analysis, interpretation, or preparation of the manuscript and approved the version presented as a preprint. All Authors contributed to the preparation and drafting of the article. Individual CRediT roles should be confirmed by the author group before journal submission.

